# Genetic overlap between Alzheimer’s disease and depression mapped onto the brain

**DOI:** 10.1101/2021.01.23.21250016

**Authors:** Jennifer Monereo Sánchez, Miranda T. Schram, Oleksandr Frei, Kevin O’Connell, Alexey A. Shadrin, Olav B. Smeland, Lars T. Westlye, Ole A. Andreassen, Tobias Kaufmann, David E.J. Linden, Dennis van der Meer

## Abstract

**Background:** Alzheimer’s disease (AD) and depression are debilitating brain disorders that are often comorbid. Shared brain mechanisms have been implicated, yet findings are inconsistent, reflecting the complexity of the underlying pathophysiology. As both disorders are (partly) heritable, characterizing their genetic overlap may provide etiological clues. While previous studies have indicated negligible genetic correlations, this study aims to expose the genetic overlap that may remain hidden due to mixed directions of effects.

**Methods:** We applied Gaussian mixture modelling, through MiXeR, and conjunctional false discovery rate (cFDR) analysis, through pleioFDR, to genome-wide association study (GWAS) summary statistics of AD (n=79,145) and depression (n=450,619). The effects of identified overlapping loci on AD and depression were tested in 403,029 participants of the UK Biobank (mean age 57.21 52.0% female), and mapped onto brain morphology in 30,699 individuals with brain MRI data.

**Results:** MiXer estimated 98 causal genetic variants overlapping between the two disorders, with 0.44 concordant directions of effects. Through pleioFDR, we identified a SNP in the *TMEM106B* gene, which was significantly associated with AD (B=-0.002, p=9.1×10^−4^) and depression (B=0.007, p=3.2×10^−9^) in the UK Biobank. This SNP was also associated with several regions of the corpus callosum volume anterior (B>0.024, p<8.6×10^−4^), third ventricle volume ventricle (B=-0.025, p=5.0×10^−6^), and inferior temporal gyrus surface area (B=0.017, p=5.3×10^−4^).

**Discussion:** Our results indicate there is substantial genetic overlap, with mixed directions of effects, between AD and depression. These findings illustrate the value of biostatistical tools that capture such overlap, providing insight into the genetic architectures of these disorders.

## Introduction

Alzheimer’s disease (AD) is a highly disabling neurodegenerative disease characterized by memory loss and a gradual cognitive, functional and behavioral decline (1). Its prevalence increases rapidly with age, affecting 13% of population at age 80, and 37% of the population at age 90 (2). Individuals with AD often have comorbid major depressive disorder (MDD), present in 22 up to 59% of cases (3,4), while MDD has an estimated lifetime prevalence of 11 to 15% in the general population (5). MDD is a heterogeneous disorder; in addition to the core symptoms of low mood, anhedonia and loss of energy, it comprises behavioral, physiological and psychological signs and symptoms that include changes in appetite, sleeping and psychomotor patterns, fatigue, lack of concentration, feelings of worthlessness or guilt, and suicidal ideation (6).

It has been long discussed whether a history of depressive symptoms is a risk factor for later development of AD, or rather an early prodromal manifestation of AD (7,8). While bidirectional effects between the two disorders is likely, there is more evidence that midlife onset depressive symptoms and/or MDD are a risk factor for AD than vice versa (9–14). Furthermore, AD patients with depressive symptoms show accelerated cognitive decline and neurodegeneration, with significantly more plaques and tangles in the hippocampus than non-depressed individuals with AD (15), while AD symptom count (16) or tau pathology (17) does not appear to contribute to the incidence or severity of depressive disorders.

Neuroimaging studies have provided scattered evidence that AD and depressive disorders share neurobiological pathways. Early stage AD is associated with atrophy of the hippocampus, para-hippocampal regions (18), and temporo-parietal cortex (19), with atrophy becoming generalized in later stages of the disease, including cortical thinning in primary motor and sensory regions (20,21). Similarly, MDD and recurrent major depression (MD) are related to smaller hippocampal volumes (22–24), amygdala and parahipocampal areas (25) as well as lower cortical thickness in medial orbitofrontal cortex, fusiform gyrus, insula, rostral and caudal anterior and posterior cingulate cortex, temporal lobe in MDD (26), many of these changes correlating positively with the duration of the disease (25). In AD patients with comorbid symptoms of depression, MRI studies have shown specifically thinner cortex in temporal and parietal areas when comparing to non-depressed AD patients (27). Conjunction analysis on the brain morphological changes overlap between AD and late-life onset depression has shown that, in addition to the previously mentioned structures, both conditions are associated with hippocampal atrophy (28). Yet, the risk of developing AD in MDD does not seem to be mediated by hippocampal or amygdala volumes (29).

Both AD and depressive disorders are heritable, with twin studies indicating 37% broad heritability for MDD (30) and 74% for AD (31). Molecular genetics studies show that both disorders have complex genetic architectures. AD has recently been characterized as oligogenic, with estimates indicating the involvement of relatively few genetic variants, in addition to the well-known, strong APOE-e4 risk variant (32,33). MDD on the other hand has been estimated to be the most polygenic of all major brain disorders (33), involving many genetic variants with small effects that explain a small amount of its heritability (34). Regardless, given the high comorbidity and indications of shared neurobiological pathways, substantial genetic overlap is to be expected, which may be leveraged to better understand these disorders (35). Indeed, several candidate gene studies have identified shared genetic risk factors (36) that implicate hypothesized shared mechanisms, such as chronic neuroinflammatory changes in the brain (37). While negligible genetic overlap between AD and MDD has been reported (38,39), substantial genetic overlap may remain hidden from measures of global genetic correlation due to mixed directions of effects. Here, we assess the genetic overlap between AD and depression across the genome through tools that capture the extent of overlap or specific loci, regardless of directions of effect. This was followed-up by analyses of the associations between shared loci and regional brain morphology in the UK Biobank (UKB) population study, providing valuable insights into their shared neurobiology.

## Materials and Methods

### Participants

We made use of data from participants of the UKB population cohort, under accession number 27412. The composition, set-up, and data gathering protocols of UKB have been extensively described elsewhere (40). We selected all individuals with White European ancestry, as determined by a combination of self-identification as ‘White British’ and similar genetic ancestry based on genetic principal components (UKB field code 22006), with good quality genetic data.

We constructed a proxy measure of AD case-control status, combining information on ICD-10 diagnoses of dementia of the participants together with parental age and parental AD status, as described previously (41). Participants with an ICD-10 diagnosis of AD (F00 and/or G30) received a score of two (n=782). All other participants received a one-unit increase for each biological parent reported to have (had) AD. Further, the contribution for each unaffected parent to the score was inversely weighted by the parent’s age/age at death, namely (100-age)/100, giving us an approximate score between 0 and 2. This approach was taken in order to account for possible late-life onset AD, i.e. to minimize the labeling of individuals that will develop AD as controls. This proxy measure has been shown to be highly genetically correlated to AD status (r_g_=0.81) (41). Participants with missing data on any of the relevant questions were excluded from these analyses (n=19,332). The final sample size was n=390,284, with a mean age of 57.33 years (SD=7.49), and 52.02% was female.

The depression phenotype utilized in this study was constructed by assigning case status to any UKB participant with an ICD10 diagnosis of depression (F32-34, F38-39), n=15,238, as well as any additional participants that answered affirmative to the question whether they had ever seen a general practitioner or psychiatrist for nerves, anxiety, tension or depression (UKB field codes 2090 and 2010), during any UKB testing visit (n=159,063). Control status was assigned to anyone who had answered “no” to these questions at all testing visits. This definition of depression is identical to the “broad depression phenotype” described by Howard in 2018 (42), based on a GWAS of depression in the UK Biobank, which reported that this definition led to the largest number of genome-wide significant hits, while still being highly genetically correlated with a GWAS using a strict clinical definition of MDD, r_g_=0.85 (42). We excluded anyone with any missing data on these questions (n=6,587). The final sample size was n=403,029, with a mean age of 57.21 years (SD=7.49), and 52.02% was female. Our sample size for the neuroimaging analyses, following pre-processing as described below and excluding individuals with brain disorders, was n=30,699. As the neuroimaging data collection took place several years after the initial data collection, this subsample had a mean age of 64.32 years (SD=7.48), and 52.06% was female.

### Genetic data pre-processing

We made use of the UKB v3 imputed data, which has undergone extensive quality control procedures as described by the UKB genetics team (43). After converting the BGEN format to PLINK binary format, we additionally carried out standard quality check procedures, including filtering out individuals with more than 10% missingness, SNPs with more than 5% missingness, SNPs with an INFO score below 0.8, and SNPs failing the Hardy-Weinberg equilibrium test at p=1*10^−9^. We further set a minor allele frequency threshold of 0.001, leaving 12,245,112 SNPs.

### Image acquisition

For the analyses involving neuroimaging data, we made use of MRI data from UKB released up to March 2020. T_1_-weighted scans were collected from four scanning sites throughout the United Kingdom, all on identically configured Siemens Skyra 3T scanners, with 32-channel receive head coils. The UKB core neuroimaging team has published extensive information on the applied scanning protocols and procedures, which we refer to for more details (44).

The T_1_-weighted scans were stored locally at the secure computing cluster of the University of Oslo. We applied the standard “recon-all -all” processing pipeline of Freesurfer v5.3, performing automated surface-based morphometry and subcortical segmentation (45,46). From the output, we extracted all commonly studied global, subcortical and cortical morphology measures, as listed in Supplementary Table 1. For each of these, we summed the left and right hemisphere measure, if applicable, leaving a total of 97 brain measures.

We excluded individuals with bad structural scan quality as indicated by an age and sex-adjusted Euler number (a measure of segmentation quality based on surface reconstruction complexity (47)) more than three standard deviations lower than the scanner site mean, or with a global brain measure more than five standard deviations from the sample mean, n=717.

### GWAS summary statistics

To investigate the genetics of AD, we made use of the phase 1 summary statistics from a recent GWAS that combined samples from the Psychiatric Genomics Consortium (PGC), the International Genomics of Alzheimer’s Project (IGAP), and the Alzheimer’s Disease Sequencing Project (ADSP) (41). The phase 1 sample of this GWAS was chosen as it did not include any UKB participants, thereby preventing sample overlap with our follow-up analyses in the UKB. The summary statistics contained 9,862,739 SNPs and was based on 24,087 late-onset AD cases and 55,058 controls with European ancestry.

For the depression phenotype of the GWAS data, we obtained the summary statistics from the Psychiatric Genomics Consortium (PGC) MDD GWAS from 2019, including the 23andMe cohort (34). The construct of depression here is based on data from cohorts with MDD as well as self-reported depression, thereby closely aligning to the measure of depression that we constructed from the UKB data. We used a version of the meta-analyzed summary statistics where the UKB sample was left out, to prevent sample overlap in downstream analyses. This version contained 15,507,882 SNPs for 121,198 individuals with depression and 329,421 controls.

For the post-GWAS analyses, we excluded the major histocompatibility complex (MHC) region (chr6:26-34MB) from both summary statistics, as well as the *APOE* locus (chr19:45 -45.8MB) from the AD GWAS, in accordance with recommendations (35). For the estimate of r_g_, we applied cross-trait linkage disequilibrium score regression (LDSR) (48). We further applied Gaussian mixture modelling, as implemented in the MiXeR tool, to the GWAS summary statistics, to estimate distributions of causal genetic variants, i.e. unobserved functional genetic variants that influence the phenotypes under investigation (49). Through MiXer, we estimated the polygenicity (the number of causal genetic variants involved) and discoverability (average effect size of the causal variants, as h^2^) of AD and depression. We further estimated the genetic overlap between AD and depression, as the number of causal variants shared regardless of direction of effects, through bivariate MiXeR.

We conducted conjunctional false discovery rate (cFDR) analysis through the pleioFDR tool using default settings (35). We set an FDR threshold of 0.05 as whole-genome significance, in accordance with recommendations (https://github.com/precimed/pleiofdr).

### Statistical analyses

All downstream analyses were carried out in R v3.6.1. In all follow-up analyses, involving UKB data, we adjusted for age, sex and the first twenty genetic principal components to control for population stratification. For the neuroimaging analyses, we additionally adjusted for scanner site, Euler number (47), and a measure-specific global estimate for the regional measures (total surface area, mean cortical thickness or intracranial volume). The latter was done to ensure that we are studying associations with regional brain morphology rather than global effects. We estimated the effective number of independent traits in our neuroimaging analyses to be 51, through spectral decomposition of the Pearson’s correlation matrix (50). We therefore set an alpha of .001 for these analyses.

Graphs were created through *ggplot2* (51), and brain maps through *ggseg* (52). The code for running pleioFDR and MiXeR is available via GitHub, https://github.com/precimed/.

## Results

### Global genetic overlap

Eighteen loci were genome-wide significant in the AD GWAS, which had an estimated SNP-based heritability, h^2^, of 0.05 (SE=0.01). The depression GWAS summary statistics contained 33 significant loci, with an h^2^ of 0.05 (SE=0.002), see Figure 1a. These numbers are in line with the results from the original GWAS studies (34,41). Using LDSC, the two disorders showed a negligible genetic correlation of −0.03 (SE=0.06, p=0.60).

**Figure 1.**
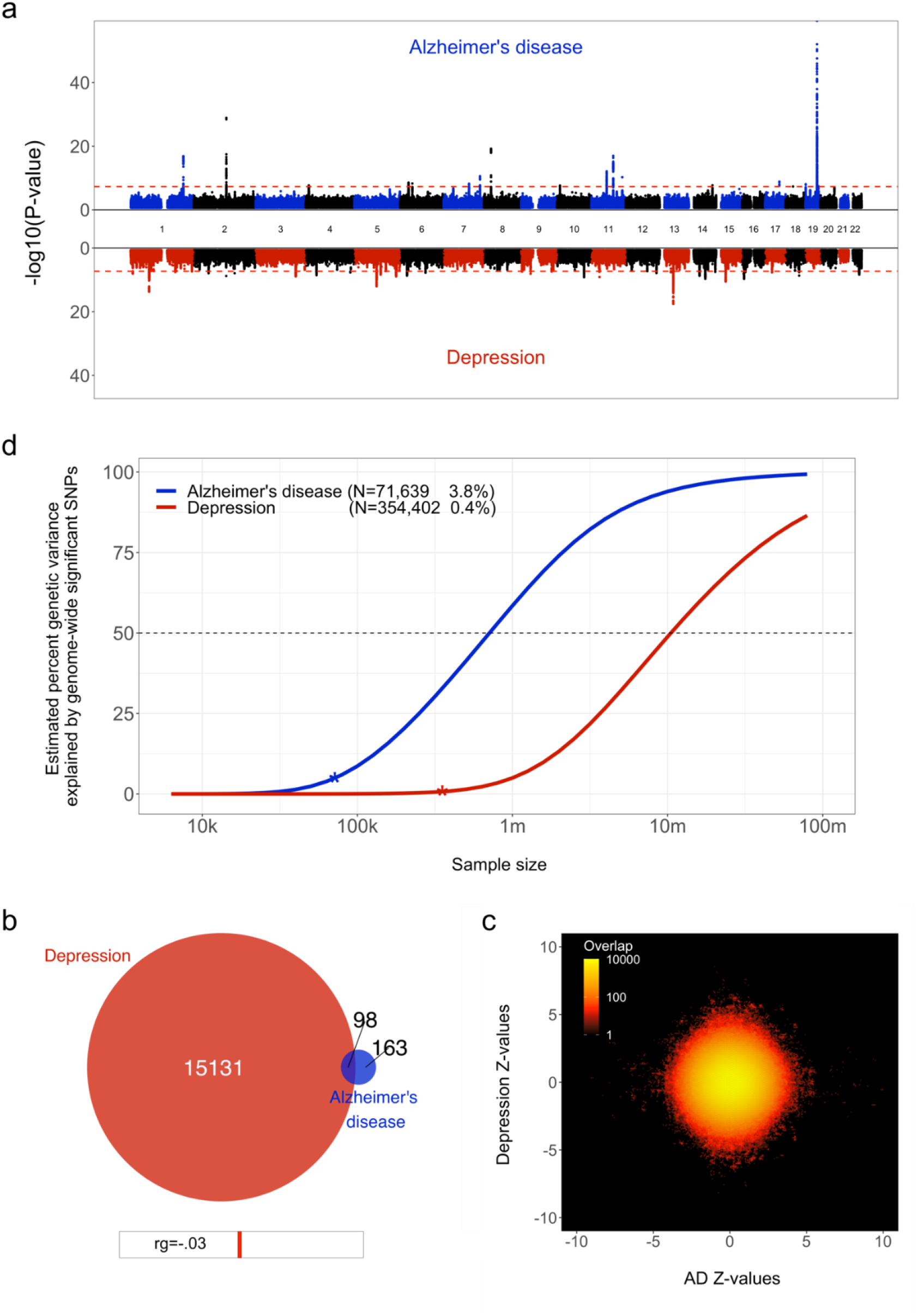
Genetic overlap between Alzheimer’s disease (AD) and depression. **a)** Miami plot, contrasting the observed -log_10_(p-values), shown on the y-axis, of each SNP for AD (top half, blue) with depression (bottom half, red). The x-axis shows the relative genomic location, grouped by chromosome, and the red dashed lines indicate the whole-genome significance threshold of 5×10^−8^. **b)** Venn diagram depicting the estimated number of causal variants shared between AD and depression and unique to either of them. Below the diagram, we show the estimated genetic correlation. **c)** Bivariate density plot, illustrating the relationship between the observed GWAS Z-values for AD (on the x-axis) and depression (on the y-axis).

Through univariate mixture modeling, we found that AD has an estimated 261 causal genetic variants, with a discoverability of 2.1×10^−4^. Depression was estimated to involve 15,228 variants, with a discoverability of 6.8×10^−6^. In other words, depression was estimated to be over fifty times more polygenic and its genetic determinants were estimated to be approximately thirty times less discoverable than AD. Expected sample sizes needed to explain half of the genetic variance for AD was 0.5 million, for depression 10 million, see Figure 1b.

Bivariate mixture modeling indicated that there were

98 causal variants overlapping between the two traits, i.e. 38% of all variants for AD and 1% of all variants for depression, see Figure 1c. The fraction of concordant directions of effects for the shared variants was 0.44. The bivariate density plot, Figure 1d, illustrates the presence of mixed directions of associations for many SNPs; some SNPs have the same direction of association for both traits, while others are positively associated with AD and negatively associated with depression or vice versa. The net result of this is a negligible negative correlation, despite a large proportion of AD’s causal variants overlapping with depression.

### Locus overlap

Through conjunctional FDR analysis, we discovered a SNP at chromosome 7, rs5011436, located at an intron of the *TMEM106B* gene, that was significantly associated with both traits. We replicated this association with both traits using UK Biobank data; for AD, we found a negative relation with the number of copies of the C allele (B=-0.002, SE=6.5×10^−4^, p=9.1×10^−4^), whereas for depression we found a positive relation (B=0.007, SE=0.001, p=3.2×10^−9^), in accordance with the directions of effects as reported in the two original GWAS.

We subsequently calculated the association of rs5011436 with cortical and subcortical brain morphology, using the neuroimaging subset of the UK Biobank. As shown in Figure 2, we found that the C allele of this SNP is significantly associated with higher volume of the posterior (B=0.035, SE=7.6×10^−3^, p=3.4×10^−6^), mid posterior (B=0.026, SE=7.5×10^−3^, p=6.6×10^−4^), and anterior (B=0.024, SE=7.3×10^−3^, p=8.6×10^−4^) sections of the corpus callosum, lower volume of the third ventricle (B=-0.025, SE=6.1×10^−3^, p=5.0×10^−6^), as well as larger area of the inferior temporal gyrus (B=0.017, SE=4.8×10^−3^, p=5.3×10^−4^).

**Figure 2.**
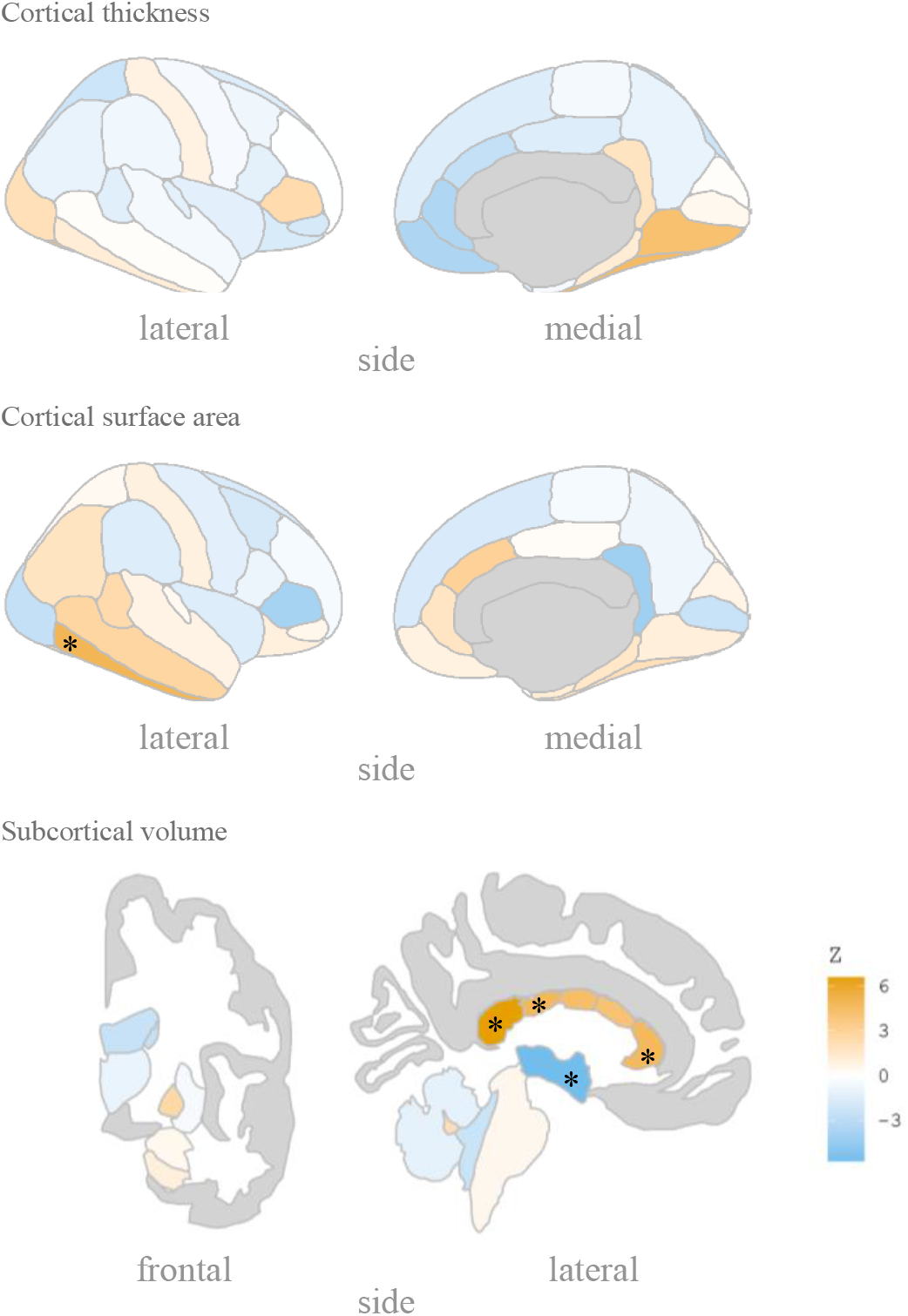
rs5011436 C allele relation to brain morphology. Brain maps showing the spatial distribution of Z scores. Legend color’s intensity shows strength in correlation. Positive correlation in orange, negative correlations in blue. Stars mark the regions that remain significant after multiple comparisons correction. Cortical thickness: no significant regions; Cortical surface area: inferior temporal gyrus; Subcortical volume: anterior, mid posterior and posterior corpus callosum, and third ventricle.

## Discussion

Here we employed state-of-the-art biostatistical tools to improve our knowledge of the genetic underpinnings of the relation between AD and depression. In line with previous reports we have identified large differences in the genetic architecture of these disorders. We add new knowledge by revealing the presence of genetic overlap between them. We further illustrated how conjunctional analysis may be used to discover specific shared genetic loci, and substantially expanded on previous efforts by mapping the effects onto the brain in order to identify neurobiological mechanisms that contribute to the relation between these disorders.

We found that many of the causal variants for AD are overlapping with depression. This partly contradicts the previously reported negligible genetic correlation between AD and MD (38), as well as the overall low genetic correlation reported between neurologic and psychiatric disorders (53). However, whereas genetic correlations rely on globally consistent directions of effects between the two traits under investigation, bivariate Gaussian mixture modelling estimates the number of causal variants that have an effect on both, regardless of directions of effects. High levels of mixed directions of effects is likely to be commonplace for complex traits such as brain disorders. This can be seen in, for instance, another psychiatric disorder like schizophrenia, which has been estimated to share virtually all causal variants with educational attainment, despite a near-zero genetic correlation (49).

The strong heterogeneity of depressive disorders is likely to contribute to the mixed directions of effects between the two traits, which appears in contradiction to the high levels of reported comorbidity with AD. The heterogeneity of the depression phenotype is evident from its wide range of signs and symptoms and the likely existence of several depression subtypes. This may explain how the extent of genetic overlap can be large for the less polygenic AD, yet very small for depression, which fits with numerous reports that depressive disorders are an important predictor of AD pathology while the opposite is less true (15–17). We speculate that some depressive disorder subtypes will be shown as genetically more concordant with AD than others, in line with indications that depressive disorders subtypes have significantly different genetic architectures (54,55). The wide range of reported levels of comorbidity with AD across studies (3,4,14,16,56) may also be due to this heterogeneity, as they differ in defining and subtyping of depression. A direct investigation of the relation of AD comorbidity and depressive disorders subtypes, coupled to neurobiological data, would be valuable. We postulate that studies using more narrow depressive disorder subtypes would find lower polygenicity and more concordant directions of effects with AD for specific subtypes.

Our use of cFDR to identify a specific locus shared by AD and depression is an example of how we may use genetics to improve our understanding of the neurobiology underlying the relation between these two disorders. The SNP rs5011436 is located in an intron of the gene *TMEM106B*, which encodes the transmembrane protein 106B. *TMEM106B* was the first genetic risk factor to be identified for fronto-temporal lobar degeneration (FTLD, 57). Since then, it has also been reported in GWAS of both AD (58) and MD (59). The protein TMEM106b is thought to regulate lysosomal function, with a role in the clearance of TPD-43 (60). Both lysosomal function and specifically TDP-43 are highly related with the pathogenesis of AD (61,62) and MD (63). *TMEM106B* expression has been shown to be downregulated in brains of individuals with AD (64), while it has been found to be upregulated in individuals with MD (65).

The evidence for involvement of *TMEM106B* in both disorders is further substantiated by our neuroimaging analyses, indicating effects on several brain regions that have been tied to both AD and depression. In particular the corpus callosum was implicated by our analyses, in line with previous neuroimaging findings on *TMEM106B* (66), with higher volume of several callosal subregions for carriers of the rs5011436 c-allele, the allele that we found to convey risk for depression and to be protective for AD. MDD is associated with abnormal cerebral lateralization, and individuals with familial MDD have been found to have significantly larger callosal volume than individuals with non-familial forms of MDD (67), while AD has been repeatedly linked to degeneration of the corpus callosum (68). Thus, specific, genetically mediated, forms of depression have been found to have opposing directions of effects on the corpus callosum than other forms of depression and AD.

Our estimates of heritability, polygenicity and discoverability highlight the complexity of the genetic architecture of both disorders. While twin studies have indicated high broad heritability of AD (31) and MD (30), we replicated previous findings of low SNP-based heritability, as captured by GWAS data (34,41,69). This possibly implicates an important role for rare variants, as well as a high degree of genetic and environmental interaction effects. Contrasting the two disorders, it is clear that AD polygenicity is relatively low, with a recent study even qualifying late-onset AD as oligogenic (32). Depressive symptoms on the other hand are highly polygenic, partly reflecting the substantial clinical heterogeneity which may capture a broad range of conditions, each likely with partly distinct genetic determinants (54). Regardless of these differences in genetic architectures, our analyses made clear that, with current approaches, GWAS sample sizes will need to reach millions of individuals to uncover a substantial fraction of the common genetic variance influencing both disorders.

Our findings once again reiterate the complexity of the genetic architectures of brain disorders, highlighting the limitations of the GWAS approach. Our power analyses suggest that, despite tremendous efforts from worldwide consortia to bring together large samples, we are only at the very beginning of uncovering the genetic determinants of AD and depression through the standard GWAS approach. Clearly, more powerful biostatistical tools are needed, ones that better match this complexity and that leverage genetic signal shared across traits of interest (35,49,70) in order to lower the required sample sizes and provide more meaningful metrics. While approaches like Gaussian mixture modeling are a step in the right direction, the current implementation does still suffer from oversimplified assumptions about the nature of the genetic architecture of brain disorders; AD is enriched for rare variants, while the MiXeR analysis focuses on common variants only. Further, low polygenicity implies a handful of large genetic effects - there is a bigger chance that the distribution of those effect sizes won’t follow a Gaussian distribution, violating model assumptions. We are developing extensions of this method that will handle such characteristics. To conclude, in this study we provided further insights into the genetic relationship between AD and depression, providing evidence of significant genetic overlap, and neuropathological effects reflected in brain morphological changes, warranting further genetic research. However, it seems that the complex relation between AD and depression will require future research to employ larger sample sizes, cleaner phenotype definitions and further improvements of biostatistical tools. It will also be important to study interaction effects between genetic variants and between genetic and environmental factors, as well as the dynamic interplay between relevant factors over the lifespan. These all will influence the underlying biological mechanisms that account for the complex relationship between these disorders. Ultimately this knowledge may provide a path towards more effective treatments, thereby reducing the enormous burden that AD and depression place on patients and their care-givers.

## Data Availability

The code for running pleioFDR and MiXeR is available via GitHub, https://github.com/precimed/.
UK biobank data is available via https://www.ukbiobank.ac.uk/

## Author contributions

J.M.S. and D.v.d.M. conceived the study; D.v.d.M., and T.K. pre-processed the data; J.M.S. and D.v.d.M. performed all analyses, with conceptual input from M.S. and D.E.J.L.; All authors contributed to interpretation of results; J.M.S. and D.v.d.M. drafted the manuscript and all authors contributed to and approved the final manuscript.

## Acknowledgements

This work was partly performed on the TSD (Tjeneste for Sensitive Data) facilities, owned by the University of Oslo, operated and developed by the TSD service group at the University of Oslo, IT-Department (USIT). We would like to thank the research participants and employees of 23andMe for making this work possible.

